# A DOUBLE-BLIND, PLACEBO-CONTROLLED, RANDOMIZED, MULTI-CENTRE, PHASE III STUDY OF MLC901 (NEUROAID II™) FOR THE TREATMENT OF COGNITIVE IMPAIRMENT AFTER MILD TRAUMATIC BRAIN INJURY

**DOI:** 10.1101/2024.08.28.24312757

**Authors:** Pavel I. Pilipenko, Anna A. Ivanova, Yulia V. Kotsiubinskaya, Vera N. Grigoryeva, Alexey Y. Khrulev, Anatoly V. Skorokhodov, Maxim M. Gavrik, Nona N. Mkrtchan, Marek Majdan, Peter Valkovic, Daria Babarova, Suzanne Barker-Collo, Kelly Jones, Valery L. Feigin

## Abstract

**Introduction:** About half of the world population will suffer from a traumatic brain injury (TBI) during their lifetime, of which about 90% of cases are mild TBI. About 15-40% of adults with TBI experience persistent cognitive deficits, and there is a lack of proven-effective treatment to facilitate cognitive recovery after mild TBI.

**Methods and analysis:** This randomized placebo-controlled multi-centre clinical trial aimed to examine the safety and efficacy of herbal supplement MLC901 (NeuroAiD II™) on cognitive functioning following mild TBI. Adults aged 18-65 years, who were 1-12-months post-mild TBI and experienced cognitive impairment, were assigned to receive either MLC901 (0.8g capsules/day) or placebo for 6 months in 7 research centres in Russia using centralized stratified permuted block randomization. The primary outcome was cognitive functioning as assessed by an online neuropsychological test (CNS Vital signs). Secondary outcomes included Rivermead Post-Concussion Symptoms Questionnaire (RPQ; neurobehavioral sequelae), Health Related Quality of Life (QOLIBRI), the Hospital Anxiety and Depression Scale (HADS), and adverse events. Assessments were completed at baseline and 3-, 6-, and 9-month follow-ups. Mixed effects models of repeated measures with intention to treat analysis were employed, with the primary outcome time-point of 6-months. A Least Square Mean Difference (LSMD) from baseline to 3-, 6-, and 9-month follow-up was calculated with 95% confidence intervals (CI).

**Results:** One hundred and eighty-two participants (mean age 40.6±14.2 in the MLC901 group and 40.1±12.0 in the Placebo group, 50% and 47.8% females, respectively) were included in the analysis. Baseline variables were comparable between groups. Multivariate mixed effects model analysis did not reveal significant improvements in complex attention (LSMD=-1.18 [95% CI -5.40; 3.03; p=0.58] and other cognitive domains at 6-months in the MLC901 group compared to the Placebo group. There were significant improvements in RPQ, QOLIBRI, anxiety and depression in the MLC901 group compared to the Placebo group at 6 and 9-months (LSMD -4.36 [-6.46; -2.26] and -4.07 [-6.22; -1.92], 4.84 [1.58; 8.10] and 3.74 [0.44; 7.03], -1.50 [-2.29; -0.71 and -0.96 [-1.84; -0.08], -1.14 [-1.92; -0.35] and -1.14 [-1.94; -0.34], respectively. No serious adverse events were reported.

**Conclusions:** The 6-month treatment with MLC901 did not result in a statistically significant difference with placebo for CNS-VS measurement of complex attention and other cognitive outcomes in individuals with mild TBI. The study showed a clinically and statistically significant improvement in all clinical scales assessed by the investigators (post-concussion symptoms, quality of life, and mood). This study showed that post-mild TBI treatment with MLC901 0.8g/day is safe.

**Trial registration:** ClinicalTrials.gov identifier NCT04861688.

## INTRODUCTION

Traumatic brain injury (TBI) is a leading cause of disability and death in young adults globally.^1,2^ It was estimated that about half of the population has suffered from a TBI at some moment in their life. There are about 50 million people experiencing new TBI annually and the burden of TBI is increasing globally.^1,3^ The annual global cost of TBI is already immense and estimated to be around 400 billion US dollars.^1^

Injury to the brain is caused by the mechanical impact of the brain onto the bony surfaces within the skull or from penetration of objects into the skull and diffuse axonal injury as a result of rotational forces as the brain moves within the skull. As a result of injury, brain cells can be damaged or die, affecting the functioning of areas that they help to control (e.g., causing neurological/cognitive deficits). The most frequent sites of cerebral contusion in closed TBI are the temporal and basal-frontal regions, both of which are associated with cognitive functioning.^4,5^

Around 90% of all TBI cases are mild TBI, and persistent cognitive deficits have been reported to occur in up to half of adults following a mild TBI and can profoundly impact a person’s day-to-day functioning, often affecting their ability to return to work, or impacting their capacity to engage in independent living.^5,6^ The most common cognitive deficits include difficulties with complex attention, executive functioning, and cognitive flexibility. After a brain injury (particularly mild injuries) many people recover spontaneously as the brain mobilizes surviving elements of the central nervous system in the damaged area to facilitate recovery. However often this spontaneous recovery process is insufficient, and people continue to experience cognitive, emotional and physical impairments.^7-9^

Despite the huge and increasing burden from mild TBI, there is still no proven-effective pharmacological treatment to improve post-TBI cognitive functioning and further research into potential new interventions is needed. Due to the complexity of the recovery processes involved after injury to the brain, there is increasing evidence that the search for a single molecule which specifically acts on a single target is not optimal.^10^ Combination therapies comprising more than one active ingredient may offer a better treatment strategy.^11^ Herbal medicine may represent a valuable resource in such a search for safe and effective therapy.

NeuroAiD II™ (MLC901) is a botanical product, derived from traditional Chinese medicine, and containing extracts from 9 herbal ingredients. It is a simplified formulation of a predecessor product, MLC601 (NeuroAiD I) which has been used in humans for many years, notably to facilitate functional recovery of patients in the post-acute phase of stroke. The herbal components of MLC601 and MLC901 are the same. In in-vitro and animal experiments, both products have been shown to protect brain cells from dying after injury, and to stimulate generation of new neural cells, connections and pathways.^12-15^ In a recent pilot placebo-controlled randomized trial conducted in New Zealand, MLC901 (NeuroAiD II™) demonstrated significant improvement in complex attention and executive functioning in individuals who had experienced some cognitive impairment after mild TBI.^16^ That pilot trial informed the design of the current study.

## METHODS

### Overview of design

The **S**afety and effic**A**cy of **M**LC901 in cognitive recovery post tra**U**matic B**RA**in **I**njury (SAMURAI) study was a phase III double-blind, placebo-controlled, randomized multicentre clinical trial in five cities/centres in Russia. The study protocol and methodology has been previously published.^17^ The primary aim of the study was to determine the cognitive benefit of using NeuroAiD II™ (MLC901) for a treatment of 6 months, compared to placebo, in adult individuals who had recently suffered a mild-TBI, and to assess its safety.

The primary efficacy outcome of the study was the measure of complex attention at month 6, one of the numerous parameters included in the battery of measures assessed by using the CNS-VS tool, an online auto-administered computerized cognitive test.^18,19^ Secondary outcomes included other parameters of the CNS-VS (executive functioning, processing speed, memory [visual and verbal] and reaction time), as well as four commonly used investigator assessment clinical outcome scales for TBI patients, namely post-concussion symptoms (as measured by Rivermead Post-Concussion Symptoms Questionnaire [RPQ]),^20^ Health Related Quality of Life After Brain Injury (QOLIBRI),^21^ anxiety and depression (as measured by the Hospital Anxiety and Depression Scale [HADS]),^22,23^ and adverse events.

The investigational product, active or placebo capsules, were identical in colour, shape and taste, and the investigators and study team remained blind to group allocation throughout the trial. An independent Data Safety and Monitoring Committee (DSMC) ensured study oversight and protection of patients’ safety, by reviewing overall data during its meetings, while remaining blinded to treatment allocation. No interim analysis was performed.

The study was conducted in accordance with the ethical principles of good clinical practice, the Declaration of Helsinki^24^ and all local regulations. SAMURAI was approved by the Ethics Committee of the Ministry of Health of the Russian Federation on 9 March 2021 (Dossier Ref#58074, meeting’s protocol #268) and all local Ethics Committees. SAMURAI is registered on ClinicalTrials.gov identifier NCT04861688. Only study participants who provided a written informed consent dated and signed at the presence of the study investigator were randomised.

### Study participants and treatments

The SAMURAI trial included 182 adults of 18-65 years of age who experienced a mild TBI in the past 1 to 12 months, had cognitive functioning difficulties as indicated by a score of >30 on the Cognitive Failures Questionnaire^25^ (consistent with criteria in the pilot study),^16^ and gave informed consent to participate in the study. Mild TBI was defined according to the WHO criteria (Glasgow Coma Score 13-15 as assessed on scene, on admission and over next 3 days; loss of consciousness for up to 30 minutes; being dazed and confused at the time of injury or post-TBI amnesia of < 24 hours duration). Exclusion criteria: (1) co-existing severe co-morbidity, including end-stage renal failure, spinal cord injury, significant substance abuse, severe liver disease, significant mental illnesses, diabetes requiring insulin injections, severe agitation, advanced cancer or other severe conditions with life expectancy of less than 5 years, as judged by the study investigator (neurologist); (2) current participation in another clinical trial within 30 days; (3) women who were pregnant or who had a positive urine pregnancy test or breast-feeding; and (4) not fluent in Russian language or have aphasia/dysphasia.

Eligible individuals were randomized using online computer system to receive either the NeuroAiD II™ (MLC901) 2 capsules (0.4g/capsule) or matched placebo orally 3 times a day for 6 months using 1:1 stratified permuted block randomization (stratified by study center, time since injury [1-3 months/4-12 months] and gender).

### Outcomes

The primary outcome measure was complex attention, as measured by an online CNS Vital signs computerized cognitive test,^18,19^ at 6 months post randomization. CNS Vital Signs is a computerized neurocognitive test battery that was developed as a routine clinical screening instrument.^18^ During CNS-VS testing (for measuring primary and secondary cognitive outcomes), after the patient completes the seven cognitive tests, the system automatically calculates scores for domains. Speed and accuracy on six tests were used to calculate the level of functioning across the following cognitive domains: complex attention, executive function, verbal memory, visual memory, processing speed, and reaction time. Raw scores were transformed to standard scores, with a mean of 100 and standard deviation (SD) of 10, based on normative data to account for age and gender effects using an integrated algorithm. Scores <90 indicate below average levels of functioning, with higher scores indicative of better cognitive functioning. The psychometric characteristics of the tests in the CNS-VS battery are very similar to the characteristics of the conventional neuropsychological tests upon which they are based.^26,27^ CNS Vital Signs has demonstrated good discriminant and concurrent validity with conventional neuropsychological tests^18^ and is sensitive to impairments across TBI severity, with evidence of good test–retest reliability.^19,28^

The Rivermead Post-Concussion Symptoms Questionnaire (RPQ)^20^ assesses neurobehavioral sequelae and consists of two subscales including the RPQ3, which includes symptoms of headaches, dizziness and nausea, and the RPQ13 comprising 13 other common symptoms such as restlessness, noise and light sensitivity, sleep disturbance, blurred vision and balance difficulties. Participants are to state the extent to which they experience each symptom in comparison to the time before accident, on a 5-point scale ranging from 0 (not experienced) to 4 (severe problem). The two subscales have revealed good test-re-test reliability and adequate external construct validity.^29,30^ A total score ranges from 0 to 64 with higher values indicating greater symptom severity.

The Quality of Life after Brain Injury measure (QOLIBRI)^21^ is an internationally validated tool to assess quality of life after brain injury.^31^ It contains two parts. The first part assesses satisfaction with health-related quality of life and is composed of six overall items and 29 items allocated to four subscales: thinking, feelings, autonomy, and social aspects. The second part, devoted to “bothered” questions, is composed of 12 items in two subscales: negative feelings and restrictions. QOLIBRI total scores <60 indicate low or impaired health-related quality of life.^32^ The QOLIBRI showed good construct validity in the TBI group.^33^

Mood was assessed by the Hospital Anxiety and Depression Scale (HADS).^34^ The scale has been widely used for assessing levels of anxiety and depression in patients with medical problems including TBI.^35^ The scale consists of 14 statements (e.g., I feel tense or ‘wound up’) that the participant is asked to rate in regards how they have been feeling in the past week, yielding separate subscale scores for anxiety and depression. Scoring for each item ranges from zero to three, with three denoting highest anxiety or depression level. The subscale scores range from 0-21 (0-7 normal, 8-10 mild, 11-14 moderate and 15-21 severe). The measure has demonstrated good test-retest reliability^36^ and good sensitivity and specificity.^37^

Adverse events were monitored at each follow-up assessment visit. An adverse event was defined as any untoward medical occurrence in a study participant that does not necessarily have a causal relationship with the treatment. Assessments of primary and secondary outcomes were completed at baseline and 1-, 3-, 6-, and 9-month follow-up.

### Statistical analyses and power calculations

Socio-demographic and clinical characteristics of the study participants at baseline were evaluated by tests of difference. For the analysis of primary and secondary endpoints, a mixed effects model (PROC MIXED)^38^ was used with adjustments for baseline and potential covariates, with the participants and sites used as the random effects. Model selection was undertaken with each outcome using standard selection heuristics. Covariates were selected based on improving the overall efficiency of the model. Regardless, baseline value of the outcome variable, age, gender, time since injury (1-3 months/4-12 months) and study center were included as covariates in the mixed effects model. Descriptive statistics were used to describe changes in complex attention and other outcome measures in the MLC901 and Placebo groups, as well as mean values and SDs for non-cognitive outcomes obtained by the method of least squares from the mixed effects model. The Wilcoxon test was used to evaluate differences in ordinal items of outcomes. Safety and tolerability were assessed by the frequency and nature of any potential adverse events recorded. Levels of adherence to the treatment regime were determined based on the self-reported number of capsules not taken. Analyses were based on the intention to treat principle.

The required sample size for the trial was based on the results of the pilot double-blind, place-controlled randomized clinical trial in New Zealand (n=78) evaluating the effect of MLC901 versus placebo using CNS Vital Signs.^16^ It was estimated that a sample of 182 participants (with 1:1 group ratio) would provide 80% statistical power (two sided α=0.05, β =0.20) to detect a clinically significant 10 points difference^28^ (SD=20) between placebo and MLC901 in the change from baseline to month 6 for the CNS Vital Signs complex attention score, assuming 30% non-compliance/lost to follow-up.

## RESULTS

The trial results were subject of an abstract presented at the World Congress of Neurology in 2023.^39^ Of the 811 individuals assessed for eligibility, 182 participants (22%) were enrolled and randomized into the study (Figure 1), with the retention rate of 98% at 9-month follow-up (one participant was lost to follow-up and three participants decided to withdraw from the study for unknown reasons). There were no significant differences between the 2 groups regarding demographic and other baseline characteristics, as shown in Table 1. The majority of mild TBIs were caused by falls (61%-62%). The median time from TBI to inclusion was 3 months. Previous history of TBI was reported in 32.6% of the MCL901 group and 40.0% of the Placebo group participants. About half of the randomized participants in both groups were females.

**Table 1.** Baseline demographic and medical characteristics of the study participants.

| **Characteristics** | **MLC901 Group**<br>**N=92** | **Placebo Group**<br>**N=90** |
| --- | --- | --- |
| Age (yrs) mean (SD) | 40.6 (14.2) | 40.1 (12.0) |
| Males (%) | 50.0 | 47.8 |
| Caucasian ethnicity (%) | 100.0 | 97.8 |
| Tertiary education or above (%) | 50.0 | 62.2 |
| Months since injury (median) | 3.0 | 2.9 |
| Full-time pre-TBI work (%) | 87.0 | 94.5 |
| Married/Partner (%) | 69.6 | 80.0 |
| Mechanism of TBI injury (%) |  |  |
| Fall | 60.9 | 62.2 |
| Motor vehicle accident | 14.1 | 13.3 |
| Violence | 9.8 | 11.1 |
| Sport trauma | 12.0 | 6.7 |
| Other | 3.3 | 6.7 |
| Prior TBI (%) | 32.6 | 40.0 |
| Other previous injuries sustained (%) | 47.8 | 55.5 |
| Baseline CNS below average (%) | 57.6 | 52.2 |

**Figure 1.**
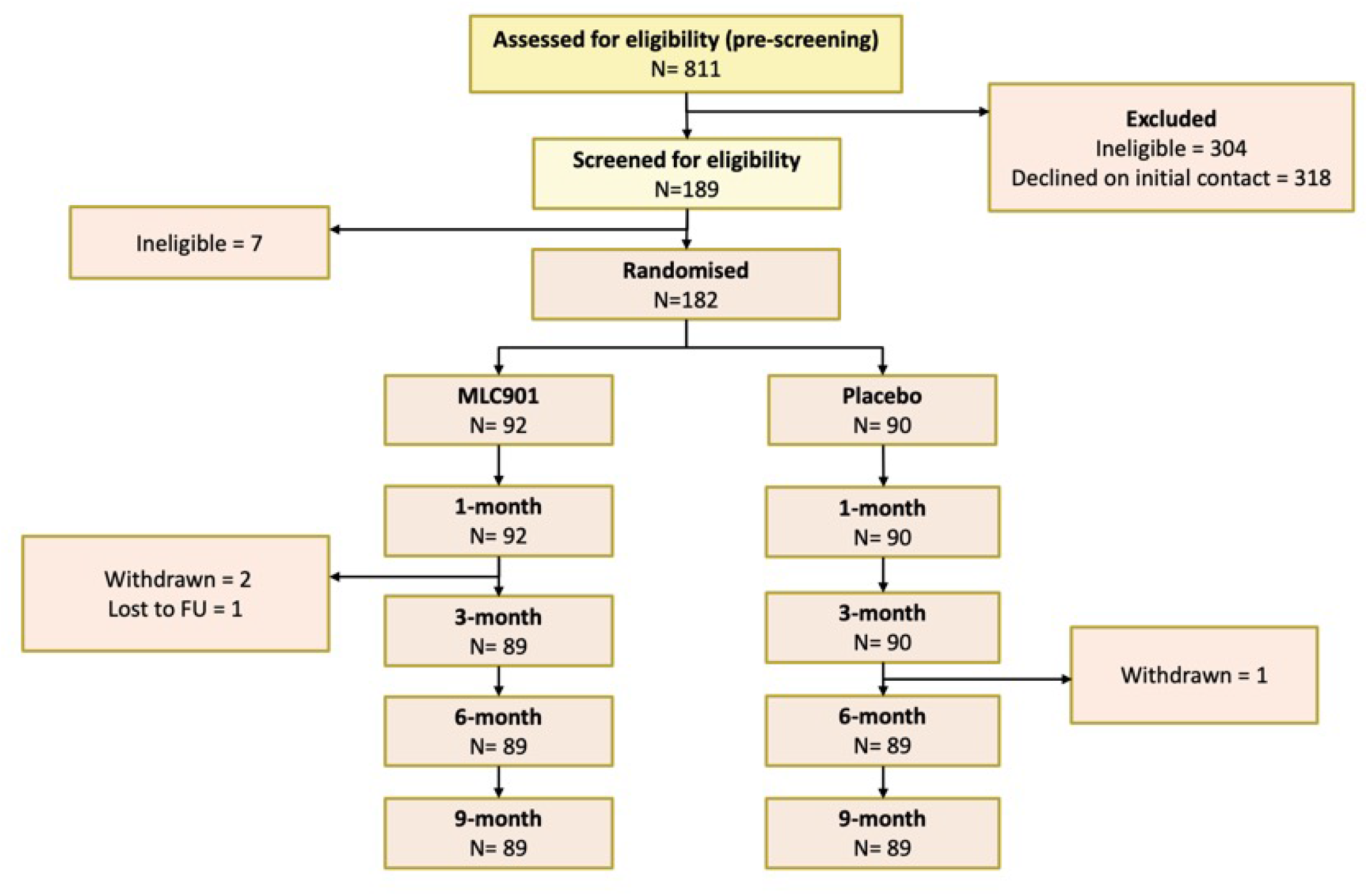
CONSORT diagram of study participants.

Twenty-seven participants in the MLC901 group (29.3%) and 34 participants in the Placebo group (37.8%) reported side effects during the 6 months post-randomization (Table 2). In regard to the most common side effect, in the intervention group, eight participants reported gastrointestinal symptoms (8.8%), as compared to three participants in the control group (3.3%). No serious adverse events (death, hospitalization, disability) were reported in either group. Overall adherence, as measured by returned pill counting, in taking the capsules varied between 99.1% (SD 2.5) for MLC901 and 98.4% (SD 10.7) for placebo at 1 month, 99.7% (SD 3.8) for MLC901 and 98.6% (SD 4.5) at 3-months and 98.9% (SD 3.9) for MLC901 and 99.5% (SD 4.3) for placebo at 6-months.

**Table 2.** Adverse effects in the MLC901 and placebo groups at 9 months post-randomization.

| Adverse events | MLC901 Group<br>N=92 | Placebo Group<br>N=90 |
| --- | --- | --- |
| Headache | 8 (8.9%) | 2 (2.2%) |
| Fatigue | 6 (6.7%) | 3 (3.3%) |
| Respiratory tract infection viral | 5 (5.6%) | 4 (4.3%) |
| COVID-19 | 4 (4.4%) | 2 (2.2%) |
| Dizziness | 3 (3.3%) | 0 |
| Nausea | 1 (1.1%) | 2 (2.2%) |
| Abdominal discomfort | 0 | 2 (2.2%) |
| Abdominal pain | 0 | 2 (2.2%) |
| Alanine aminotransferase increased | 1 (1.1%) | 1 (1.1%) |
| Aspartate aminotransferase increased | 1 (1.1%) | 1 (1.1%) |
| Bronchitis | 1 (1.1%) | 1 (1.1%) |
| Diarrhoea | 0 | 2 (2.2%) |
| Nasopharyngitis | 1 (1.1%) | 1 (1.1%) |
| Respiratory tract infection | 1 (1.1%) | 1 (1.1%) |
| Abdominal distension | 1 (1.1%) | 0 |
| Anterograde amnesia | 0 | 1 (1.1%) |
| Asthenia | 0 | 1 (1.1%) |
| Biliary colic | 0 | 1 (1.1%) |
| Cholelithiasis | 0 | 1 (1.1%) |
| Decreased appetite | 0 | 1 (1.1%) |
| Dermatitis acneiform | 1 (1.1%) | 0 |
| Disturbance in attention | 1 (1.1%) | 0 |
| Eye injury | 1 (1.1%) | 0 |
| Haematuria | 0 | 1 (1.1%) |
| Numbness in arms or legs | 0 | 1 (1.1%) |
| Hyposmia | 0 | 1 (1.1%) |
| Hypothyroidism | 0 | 1 (1.1%) |
| Increased appetite | 0 | 1 (1.1%) |
| Insomnia | 0 | 1 (1.1%) |
| Irritable bowel syndrome | 1 (1.1%) | 0 |
| Laryngitis | 1 (1.1%) | 0 |
| Memory impairment | 1 (1.1%) | 0 |
| Osteoarthritis | 0 | 1 (1.1%) |
| Otitis media acute | 1 (1.1%) | 0 |
| Photophobia | 1 (1.1%) | 0 |
| Rhinitis | 1 (1.1%) | 0 |
| Rhinitis allergic | 0 | 1 (1.1%) |
| Sleep disorder | 0 | 1 (1.1%) |
| Spinal pain | 0 | 1 (1.1%) |
| Taste disorder | 0 | 1 (1.1%) |
| Tinnitus | 1 (1.1%) | 0 |
| Tonsillitis | 1 (1.1%) | 0 |
| Tremor | 1 (1.1%) | 0 |
| Vertigo | 0 | 1 (1.1%) |
| Viral infection | 0 | 1 (1.1%) |
| Vision blurred | 1 (1.1%) | 0 |

The assessment of cognitive impairment made by the patient by using the CNS-VS tool did not reveal a statistically significant difference between placebo and MLC901 (Table 3). This was the case for the primary outcome (change from month 6 to baseline in complex attention (LSMD=-1.18 [95% CI -5.40; 3.03], p=0.58), as well as for the other CNS-VS derived cognitive parameters (executive functioning, visual memory, verbal memory, processing speed, or reaction time).

**Table 3.**
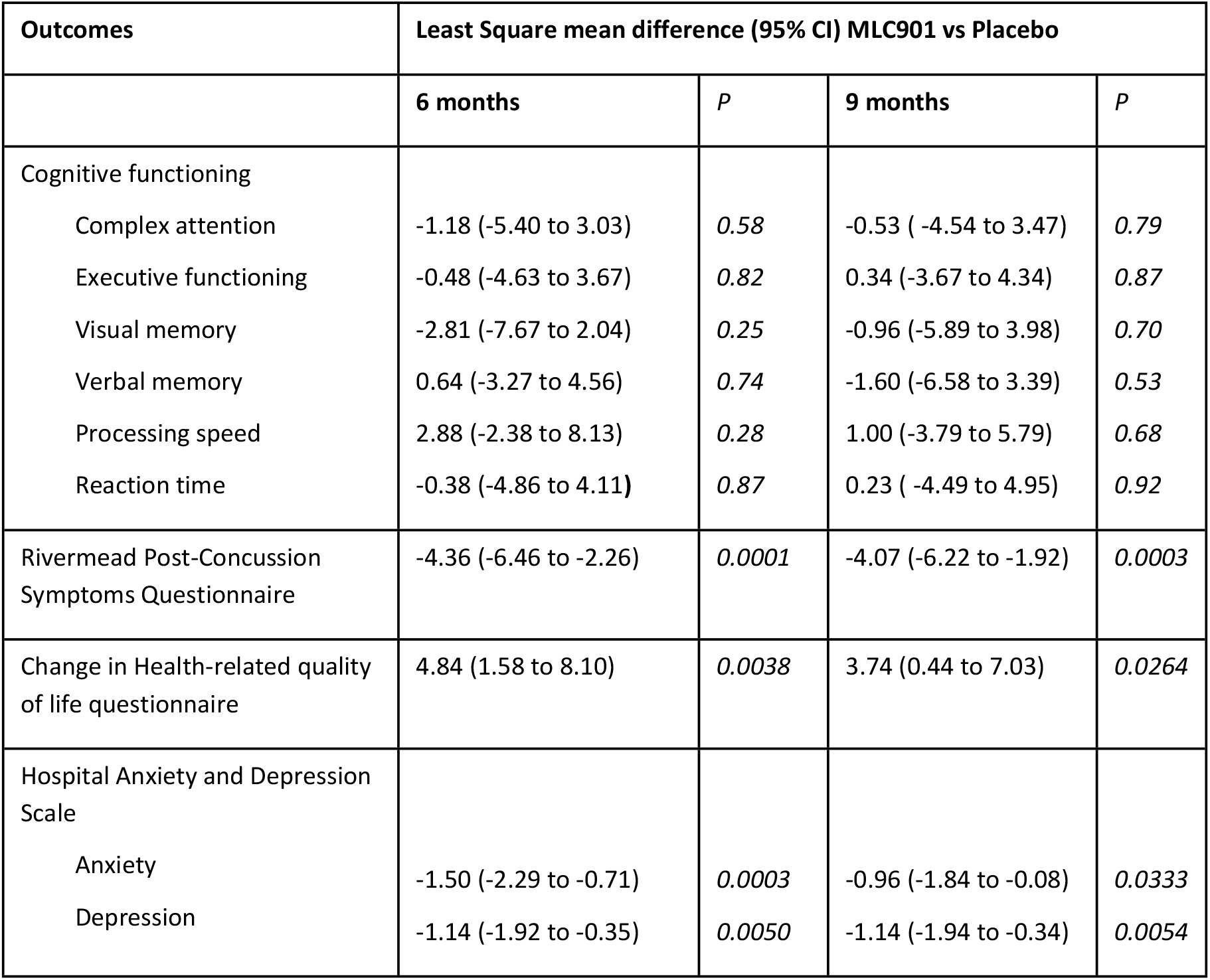
Least square mean difference, with 95% CI, in primary and secondary outcomes between MLC901 group and Placebo group at 6 and 9 months after randomisation.

Participants randomized to receive MLC901 had statistically and clinically significant improvements in post-concussion symptoms, quality of life, anxiety and depression at 6-and 9-months post-randomization compared to participants randomized to receive Placebo (Table 3; Figures 1-4). Importantly, the improvement continued up to 9 months of follow-up although active treatment was stopped at 6-months post-randomization. By the 9month follow-up the Rivermead Post-Concussion Symptoms score improved in the MCL901 group by 47% (95% CI 41% - 53%), while in the Placebo group it improved by only 29% (95% CI 22% - 35%), quality of life improved in the MLC901 group by 22% (95% CI 18% - 26%) and only 14% (96% CI 11% - 18%) in the Placebo group, anxiety score improved in MLC901 group by 49% (95% CI 41% - 57%) and only 42% (95% CI 33% - 51%) in the Placebo group, and depression score reduced by 48% (95% CI 39% - 57%) in the MLC901 group and only 31% (95% CI 22% - 40%) in the Placebo group. Pre-determined subgroup sensitivity analysis with exclusion of missing values, outliers by age, sex at birth, history of cancer, previous TBI, and time since mild TBI event onset did not reveal any significant difference in cognitive outcomes between the groups (these results are not shown).

## DISCUSSION

To the best of our knowledge this placebo-controlled randomized full-scale trial is the first to determine efficacy of MLC901 (2 capsules 3 times [0.8 g a day] a day for 6 months) on cognitive functioning and post-mild TBI symptoms compared to a matched placebo in adults 16-65 years old with mild TBI. The trial failed to demonstrate a statistically significant difference in the CNS-VS computerized assessment of complex attention (Figure 1) and other CNS-VS measures between the MLC901 group and the Placebo group (Supplement Table 1). However, the assessment of post-TBI symptoms using standard clinical non-cognitive outcome scales used in this setting has shown statistically significant difference at both 6 and 9 months in favour of MLC901. These findings were consistent for all clinical outcome scales used in the trial.

Up to 20% of people with mild TBI show persistent post-concussion symptoms,^40^ about one third of people with mild TBI experience reduced quality of life,^31,41^ and many of them suffer from anxiety or depression.^42^ Analysis of common post-TBI somatic symptoms in our trial measured by the Rivermead Post-Concussion Questionnaire (headaches, dizziness, noise sensitivity, sleep disturbance, fatigue, irritability, depression, poor memory, poor concentration, blurred vision, light sensitivity, double vision, restlessness),^20^ quality of life (as measured by the Quality of Life after Brain Injury measure - QOLIBRI),^21^ anxiety and depression (as measured by the Hospital Anxiety and Depression Scale)^22^ showed clinically and statistically significant improvement in the MLC901 group compared to the Placebo group: LSMD=-4.36 (−6.46 to -2.26); 4.84 (1.58 to 8.10), and -1.50 (−2.29 to -0.71) and -1.14 (− 1.92 to -0.35), respectively (Figures 2-5). Importantly, the improvements tended to continue and were even enhanced at 9-months of follow-up while the active treatment was stopped at 6 months post-randomization, suggesting that the treatment somehow corrected the pathophysiological mechanisms responsible for these symptoms and triggered a self-recovering process. It remains unclear how long this recovery process might continue in both groups.

**Figure 2.**
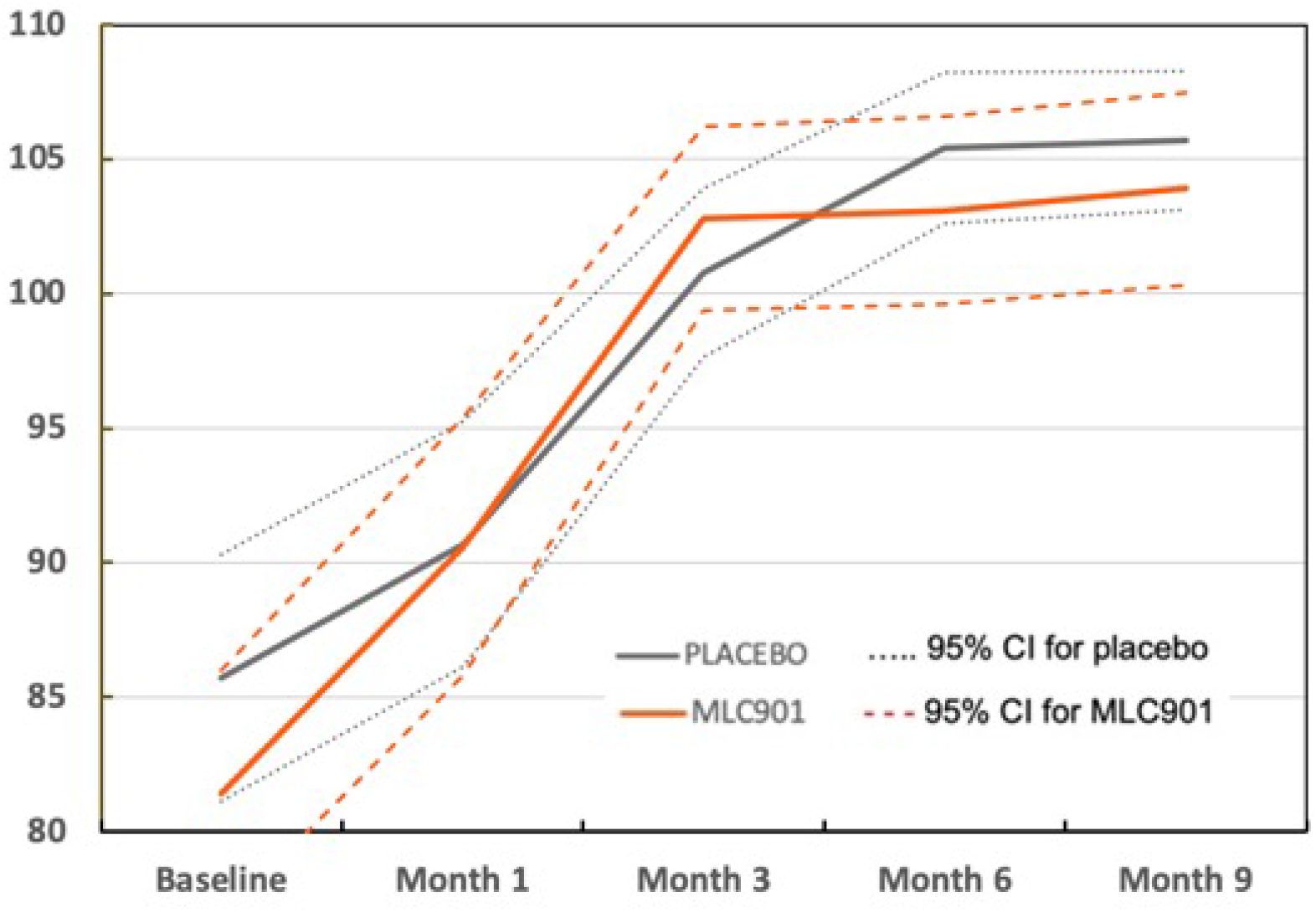
Changes in complex attention by CNS Vital Signs across the follow-up time points in MLC901 group compared to Placebo group across 9 months of follow-up period. Least square mean difference at 6 months = -1.18 (95% CI -5.40 to 3.03); p=0.58

**Figure 3.**
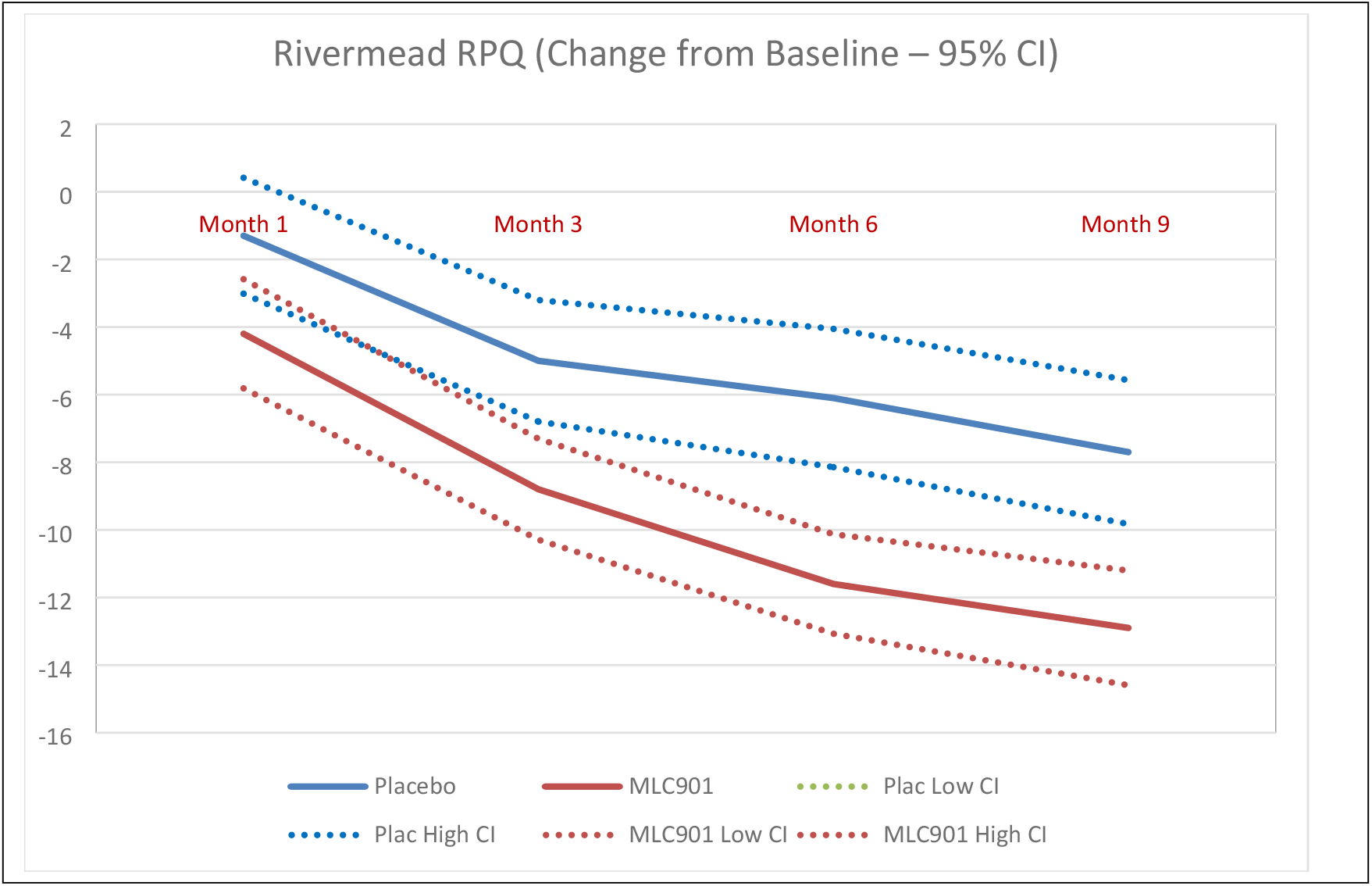
Changes in the Rivermead Scale across the follow-up time points in MLC901 group compared to Placebo group across 9 months of follow-up period. Least square mean difference at 6 months = 4.36 (−6.46 to -2.26); p = 0.0001

**Figure 4.**
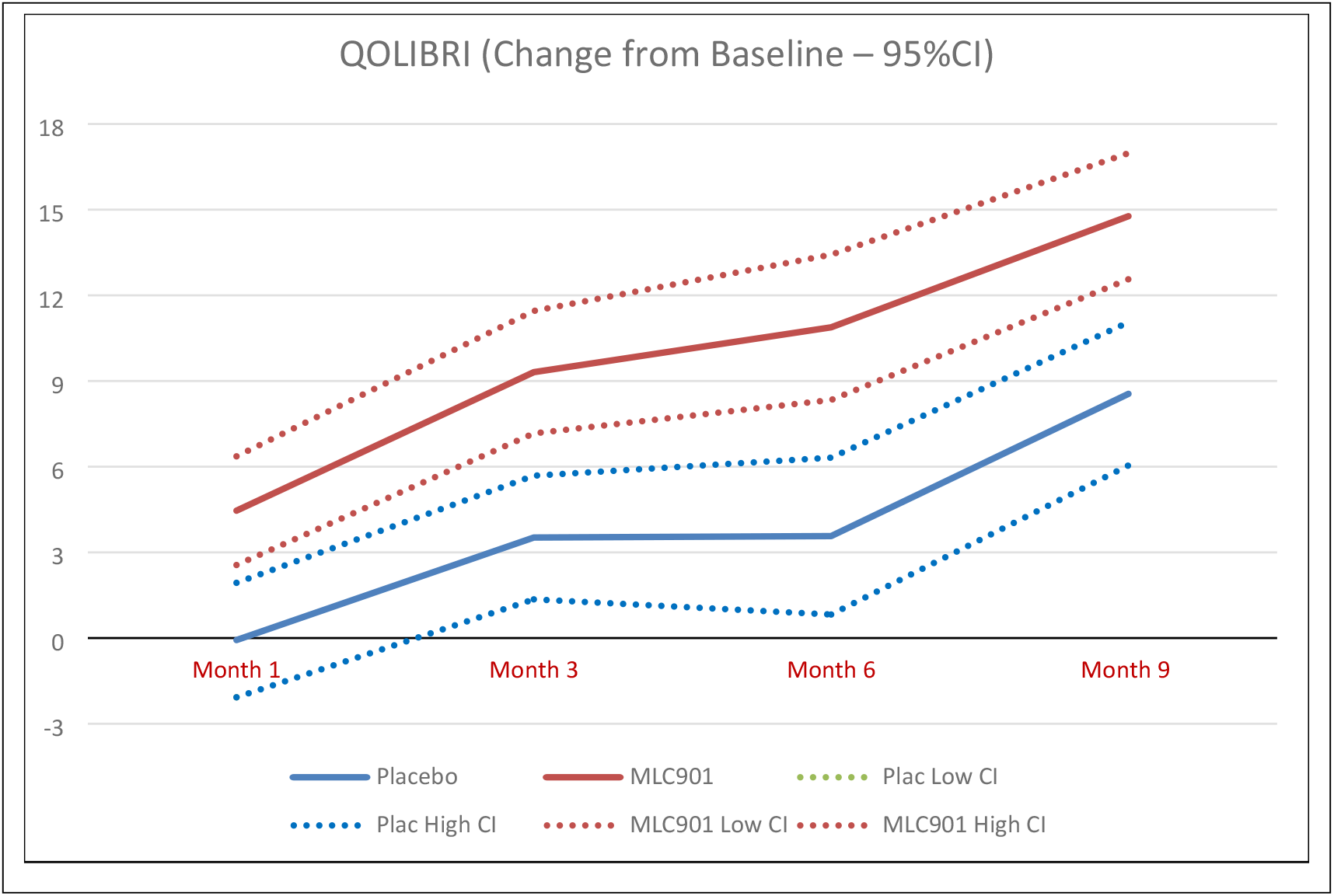
Changes in the Quality of Life after Brain Injury measured across the follow-up time points in MLC901 group compared to Placebo group across 9 months of follow-up period. Least square mean difference at 6 months = 4.84 (1.58 to 8.10); p = 0.0038

**Figure 5.**
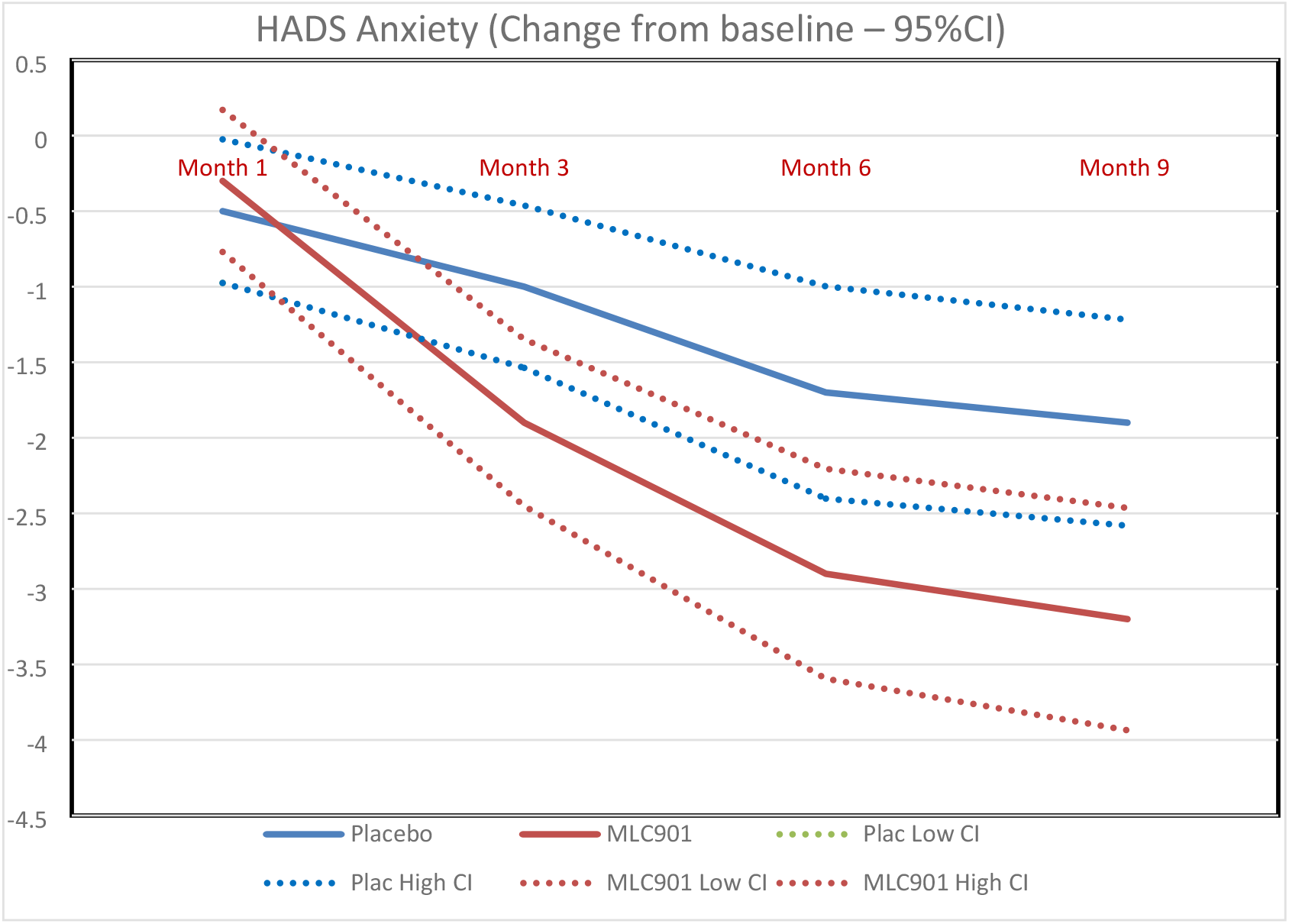
Changes in the anxiety score, as measured by the Hospital Anxiety and Depression Scale, across the follow-up time points in MLC901 group compared to Placebo group across 9 months of follow-up period. Least square mean difference at 6 months = -1.50 (−2.29 to -0.71); p = 0.0003

**Figure 6.**
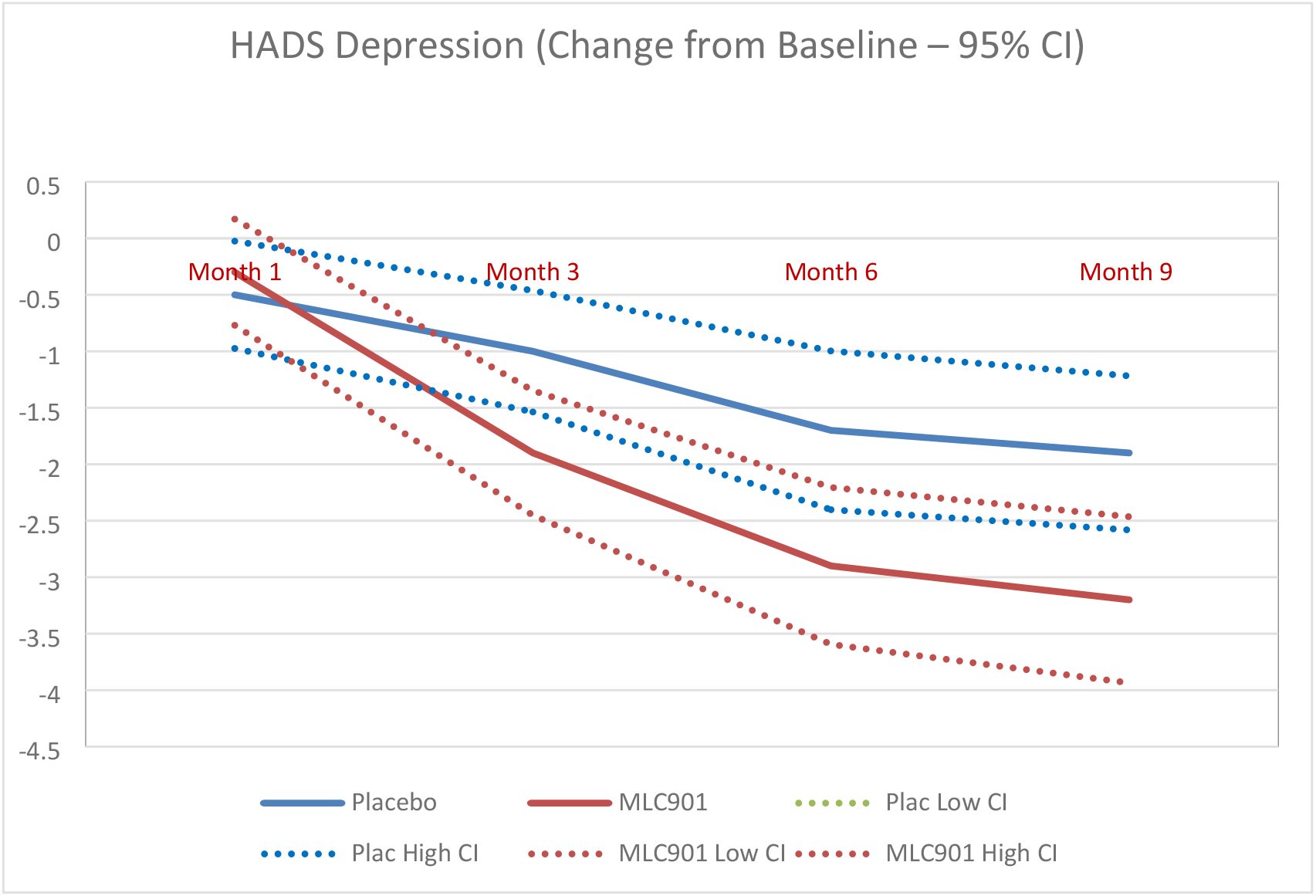
Changes in the depression score, as measured by the Hospital Anxiety and Depression Scale, across the follow-up time points in MLC901 group compared to Placebo group across 9 months of follow-up period. Least square mean difference at 6 months = -1.14 (−1.92 to -0.35); p= 0.0050

The trial also showed an excellent safety and tolerability profile. The number of side effects (none of them being considered as serious adverse event [SAE]) was low, and they were more often observed in the placebo group than in MLC901 group (Table 2). None of the side effects required discontinuation of the treatment. Moverover, adherence to the trial medications across the 6 months of the treatment was close to 100%.

Comparison of results of this trial with the previous pilot placebo-controlled trial of MLC901 in New Zealand^16^ showed that, contrary to the finding of the earlier study, the full-scale trial did not show a statistically significant improvement in complex attention, as measured by the computerized CNS-VS tool. However, the CNS-VS tool is relatively complex to use, and there is very little experience of its use in Russia. It is therefore possible that some questions and instructions used in performing the tests were not done properly in the study in Russia.

This assumption is 2orted by the observation of statistically significant improvement in some important items of the commonly used and well validated in Russia Rivermead Post-Concussion Questionnaire,^43^ specifically forgetfulness/poor memory (P=0.008 at 6 months and P=0.006 at 9 months of follow-up) and concentration function (P=0.002 at 6 months and P=0.049 at 9 months of follow-up) in the MLC901 group compared to the Placebo group. A cognitive sub-item of the Rivermead Post-Concussion Questionnaire, the speed of processing information (question “taking longer to think”), also demonstrated a statistically significant improvement) in the MLC901 group compared to the Placebo group (P=0.009; for details see Supplement Table 2). This is a particularly important clinical outcome, because speed processing is shown to be one of the most important indicators of diffuse axonal injury and TBI severity.^44,45^ The lack of statistical difference obtained for the CNS-VS endpoints in mild TBI subjects may also be related to the insufficient ability of the CNS VS to detect subtle changes in cognition 6 to 9 months after a mild TBI, as there is evidence that long after a mild TBI, high-functioning young adults invoke a strategy of delaying their identification of targets in order to maintain, and facilitate, accuracy on cognitively demanding tasks.^46^

However, both trials showed no statistically significant effect of MCL901 on other cognitive domains of CNS Vital Signs. Although both trials were not statistically powered to reliably assess effect of MLC901 on post-concussion symptoms, quality of life and mood, the direction of the treatment effect on these secondary outcomes in both trials and highly statistically significant and consistent treatment effect of MLC901 on these secondary outcomes in the full-scale trial across all follow-up time-points (including 3 months after cessation of the treatment) may suggest that this treatment effect may not be just by chance. Sensitivity analysis in various age and sex groups, different time points after TBI onset and randomization, and exclusion of outliers did not significantly change the positive treatment effect of MLC901 on post-concussion symptoms, quality of life, and mood, thus supporting the robustness of the results. These observations deserve further investigation.

Although our trial met most of the criteria for ‘gold standard’ trials (placebo-controlled, double-blinded, proper quality randomization technique, high generalizability of the trial results [wide inclusion and narrow exclusion criteria, heterogeneous study population with multi-centre-settings], fully-statistically powered with a relatively large sample size, and very low [<10%] attrition rate), it was not free from limitations. The main limitation of the trial was lack of use and absence of validation of the computerized CNS-Vital Signs test in Russia. CNS-VS is also only a cognitive screening tool, not validated for treatment evaluation. CNS-Vital Signs data was also missing for a small number of participants in both groups (5 in MLC901 group and 5 in Placebo group) and outliers in regard to missing data in some of the recruitment sites (i.e., the majority of missing data 8 [80%] came from a single site). However, sensitivity analyses with various assumptions, including exclusion of outliers, did not significantly influence the neurocognitive test results. We also did not record adverse effects right after cessation of the experimental treatment (6-month post-randomisation), but only at 9 months post-randomization.

In summary, although the trial was negative for the primary outcome, it was highly positive for other important clinical post-TBI outcomes. Given the perfect safety and tolerability of MLC901 and absence of other proven effective medications for treatment post-mild TBI symptoms, mood and health-related quality of life, a trial treatment with MLC901 in selected mild TBI adults may be worth considering for reducing post-concussion symptoms, anxiety/depression and improving health-related quality of life.

## Data Availability

All relevant data are within the manuscript and its Supporting Information files.

## Authors contributions

Study conception and design was primarily performed by Prof. Pavel I. Pilipenko, MD, PhD and Prof. Valery L. Feigin, MD, PhD, who both drafted the first version of the manuscript. Study conduct, including screening of patients, obtaining written informed consent, follow-up and assessment of safety and efficacy and data collection was done by the study Lead Principal Investigator Prof. Pavel I. Pilipenko, MD, PhD; and teams lead by Principal Investigators Dr Anna A. Ivanova, MD; Dr Yulia V. Kotsiubinskaya, MD, PhD; Prof. Vera N. Grigoryeva, MD, PhD; Dr Alexey Y. Khrulev, MD, PhD; Dr Anatoly V. Skorokhodov, MD; Dr Maxim M. Gavrik, MD, Dr. Nona N. Mkrtchan, MD. Monitoring, Data management and statistical analysis were done by an independent CRO “Atlant Clinical”. The Independent Data Monitoring Committee (IDMC) was chaired by Prof. Marek Majdan, PhD, with committee members being Prof. Peter Valkovic, PhD and Dr Daria Babarova, PhD. Overall scientific oversight for the study was provided by Prof. Suzanne Barker-Collo, PhD; A/Prof. Kelly Jones, PhD; Prof. Valery L. Feigin, MD, PhD. All authors have reviewed, provided an intellectual input into the content and approved the final manuscript.

## Disclosures

The study was funded by Moleac Pte Ltd, Singapore who manufactures the MLC901 supplement. The authors declare no financial or other conflicts of interest.

## ACKOWLEDGEMENTS

We would like to acknowledge the financial support of Moleac Pte Ltd and thank the participants for their time and interest in taking part in this study.

## Notes

### Competing Interest Statement

The authors have declared no competing interest.

### Clinical Trial

NCT04861688

### Funding Statement

The author(s) received no specific funding for this work.

### Author Declarations

Ethics Committee of the Ministry of Health of the Russian Federation on 9 March 2021 (Dossier Ref#58074, meeting’s protocol #268) and all local Ethics Committees

## References

1. Maas AIR, Menon DK, Adelson PD, Andelic N, Bell MJ, Belli A, … Zumbo F. Traumatic brain injury: integrated approaches to improve prevention, clinical care, and research. The Lancet Neurology. 2017;16:987–1048. doi: 10.1016/S1474-4422(17)30371-X

2. Langlois JA, Rutland-Brown W, Wald MM. The epidemiology and impact of traumatic brain injury: a brief overview. Journal of Head Trauma Rehabilitation. 2006;21:375–378.

3. Feigin VF, Theadom A, Barker-Collo SL, Starkey N, McPherson K, Kahan M, … Group ftBS. Incidence of traumatic brain injury in New Zealand: a population-based study. The Lancet Neurology. 2013;12:53–64.

4. Halliwell B. Reactive oxygen species and the central nervous system. Journal of Neurochemistry. 1992;59:1609–1623.

5. McMahon P, Hricik A, Yue JK, Puccio AM, Inoue T, Lingsma HF, … Vassar MJ. Symptomatology and Functional Outcome in Mild Traumatic Brain Injury: Results from the Prospective TRACK-TBI Study. J Neurotrauma. 2013;31 Epub ahead of print.

6. Theadom A, Parag V, Dowell T, McPherson K, Starkey N, Barker-Collo S, … Group BR. Persistent problems 1 year after mild traumatic brain injury: a longitudinal population study in New Zealand. Br J Gen Pract. 2016;66:e16–23. doi: 10.3399/bjgp16X683161

7. Iverson GL. Outcome from mild traumatic brain injury. Current opinion in psychiatry. 2005;18:301–317. doi: 10.1097/01.yco.0000165601.29047.ae

8. Lannsjo M, af Geijerstam JL, Johansson U, Bring J, Borg J. Prevalence and structure of symptoms at 3 months after mild traumatic brain injury in a national cohort. Brain Inj. 2009;23:213–219.

9. Zumstein MA, Moser M, Mottini M, Ott SR, Sadowski-Cron C, Radanov BP, … Exadaktylos A. Long-term outcome in patients with mild traumatic brain injury: a prospective observational study. Journal of Trauma. 2011;7:120–127.

10. Lynch, D.G.; Narayan, R.K.; Li, C. Multi-Mechanistic Approaches to the Treatment of Traumatic Brain Injury: A Review. J. Clin. Med. 2023, 12, 2179. https://www.preprints.org/manuscript/202302.0471/v1.

11. Kawata K, Rettke DJ, Thompson C, Mannix R, Bazarian JJ, Datta D. Effectiveness of biomedical interventions on the chronic stage of traumatic brain injury: a systematic review of randomized controlled trials. Front Neurol. 2024;15:1321239. doi: 10.3389/fneur.2024.1321239

12. Tsai MC, Chang CP, Peng SW, Jhuang KS, Fang YH, Lin MT, … Tsao TC. Therapeutic efficacy of Neuro AiD™ (MLC 601), a traditional Chinese medicine, in experimental traumatic brain injury. J Neuroimmune Pharmacol. 2015;10:45–54. doi: 10.1007/s11481-014-9570-0

13. Ranuh I, Sari GM, Utomo B, Suroto NS, Fauzi AA. Systematic Review and Meta-Analysis of the Efficacy of MLC901 (NeuroAiD II(TM)) for Acute Ischemic Brain Injury in Animal Models. J Evid Based Integr Med. 2021;26:2515690x211039219. doi: 10.1177/2515690x211039219

14. Heurteaux C, Gandin C, Borsotto M, Widmann C, Brau F, Lhuillier M, … Lazdunski M. Neuroprotective and neuroproliferative activities of NeuroAid (MLC601, MLC901), a Chinese medicine, in vitro and in vivo. Neuropharmacology. 2010;58:987–1001. doi: 10.1016/j.neuropharm.2010.01.001

15. Quintard H, Borsotto M, Veyssiere J, Gandin C, Labbal F, Widmann C, … Heurteaux C. MLC901, a traditional Chinese medicine protects the brain against global ischemia. Neuropharmacology. 2011;61:622–631. doi: 10.1016/j.neuropharm.2011.05.003

16. Theadom A, Barker-Collo S, Jones KM, Parmar P, Bhattacharjee R, Feigin VL. MLC901 (NeuroAiD II) for cognition after traumatic brain injury: a pilot randomized clinical trial. Eur J Neurol. 2018;25:1055–e1082. doi: 10.1111/ene.13653

17. Pilipenko P, Ivanova AA, Kotsiubinskaya YV, Feigin V, Majdan M, Grigoryeva VN, … Khrulev AY. Randomised, double-blind, placebo-controlled study investigating Safety and efficAcy of MLC901 in post-traUmatic bRAin Injury: the SAMURAI study protocol. BMJ Open. 2022;12:e059167. doi: 10.1136/bmjopen-2021-059167

18. Gualtieri CT, Johnson LG. Reliability and validity of a computerized neurocognitive test battery, CNS Vital Signs. Archives of Clinical Neuropsychology. 2006;21:623–643.

19. Gualtieri CT, Johnson LG, Benedict KB. Psychometric and clinical properties of a new, computerized neurocognitive asessment battery. In: American Neuropsychiatric Association Annual Meeting. Bal Harbor, FL; 2004.

20. King NS, Crawford S, Wenden FJ, Moss NE, Wade DT. The Rivermead Post Concussion Symptoms Questionnaire: a measure of symptoms commonly experienced after head injury and its reliability. Journal of Neurology. 1995;242:587–592.

21. Von Steinbuechel N, Petersen C, Bullinger M. Assessment of health-related quality of life in persons after traumatic brain injury - Development of the Qolibri, a specific measure. In; 2005:43–49.

22. Zigmond AS, Snaith RP. The Hospital Anxiety and Depression Scale. Acta Psychiatrica Scandinavica. 1983;67:361–370.

23. Bjelland I, Dahl AA, Haug TT, Neckelmann D. The validity of the Hospital Anxiety and Depression Scale. An updated literature review. Journal of Psychosomatic Research. 2002;52:69–77.

24. International Council for Harmonisation of Technical Requirements for Pharmaceuticals for Human Use (ICH). ICH Harmonised Guideline Integrated Addendum to ICH E6(R1): Guideline for Good Clinical Practice E6(R2); 2016. chrome-extension://efaidnbmnnnibpcajpcglclefindmkaj/https://database.ich.org/sites/default/files/E6_R2_Addendum.pdf Accessed 30 October 2023.

25. Broadbent DE, Cooper PF, FitzGerald P, Parkes KR. The Cognitive Failures Questionnaire (CFQ) and its correlates. British Journal of Clinical Psychology. 1982;21:1–16.

26. Gualtieri CT, Johnson LG. Reliability and validity of a computerized neurocognitive test battery, CNS Vital Signs. Arch Clin Neuropsychol. 2006;21:623–643. doi: 10.1016/j.acn.2006.05.007

27. Papathanasiou A, Messinis L, Georgiou VL, Papathanasopoulos P. Cognitive impairment in relapsing remitting and secondary progressive multiple sclerosis patients: efficacy of a computerized cognitive screening battery. ISRN Neurol. 2014;2014:151379. doi: 10.1155/2014/151379

28. Gualtieri CT, Johnson LG. A computerized test battery sensitive to mild and severe brain injury. Medscape journal of medicine. 2008;10:90.

29. Eyres S, Carey A, Gilworth G, Neumann V, Tennant A. Construct validity and reliability of the Rivermead Post-Concussion Symptoms Questionnaire. Clin Rehabil. 2005;19:878–887. doi: 10.1191/0269215505cr905oa

30. King NS, Crawford S, Wenden FJ, Moss NE, Wade DT. The Rivermead Post Concussion Symptoms Questionnaire: a measure of symptoms commonly experienced after head injury and its reliability. J Neurol. 1995;242:587–592. doi: 10.1007/BF00868811

31. Rauen K, Späni CB, Tartaglia MC, Ferretti MT, Reichelt L, Probst P, … Plesnila N. Quality of life after traumatic brain injury: a cross-sectional analysis uncovers age- and sex-related differences over the adult life span. GeroScience. 2021;43:263–278. doi: 10.1007/s11357-020-00273-2

32. Wilson L, Marsden-Loftus I, Koskinen S, Bakx W, Bullinger M, Formisano R, … Truelle JL. Interpreting Quality of Life after Brain Injury Scores: Cross-Walk with the Short Form-36. J Neurotrauma. 2017;34:59–65. doi: 10.1089/neu.2015.4287

33. von Steinbuchel N, Wilson L, Gibbons H, Hawthorne G, Hofer S, Schmidt S, … Force QT. Quality of Life after Brain Injury (QOLIBRI): scale validity and correlates of quality of life. J Neurotrauma. 2010;27:1157–1165. doi: 10.1089/neu.2009.1077

34. Zigmond AS, Snaith RP. The hospital anxiety and depression scale. Acta Psychiatr Scand. 1983;67:361–370. doi: 10.1111/j.1600-0447.1983.tb09716.x

35. Whelan-Goodinson R, Ponsford J, Schönberger M. Validity of the Hospital Anxiety and Depression Scale to assess depression and anxiety following traumatic brain injury as compared with the Structured Clinical Interview for DSM-IV. J Affect Disord. 2009;114:94–102. doi: 10.1016/j.jad.2008.06.007

36. Herrmann C. International experiences with the Hospital Anxiety and Depression Scale--a review of validation data and clinical results. J Psychosom Res. 1997;42:17–41. doi: 10.1016/s0022-3999(96)00216-4

37. Bjelland I, Dahl AA, Haug TT, Neckelmann D. The validity of the Hospital Anxiety and Depression Scale. An updated literature review. J Psychosom Res. 2002;52:69–77. doi: 10.1016/s0022-3999(01)00296-3

38. Thiébaut R, Jacqmin-Gadda H, Chêne G, Leport C, Commenges D. Bivariate linear mixed models using SAS proc MIXED. Computer Methods and Programs in Biomedicine. 2002;69:249–256. doi: 10.1016/S0169-2607(02)00017-2

39. Pilipenko PI, Ivanova AA, Kotsiubinskaya YV, Grigoryeva VN, Khrulev AY, Skorokhodov AV, … Feigin V. A double-blind, placebo-controlled, randomised, multi-centre, phase III study of MLC901 (neuroaid) for the treatment of cognitive impairment after mild traumatic brain injury. Journal of the Neurological Sciences. 2023;455:121095. doi: 10.1016/j.jns.2023.121095

40. Heslot C, Azouvi P, Perdrieau V, Granger A, Lefèvre-Dognin C, Cogné M. A Systematic Review of Treatments of Post-Concussion Symptoms. J Clin Med. 2022;11. doi: 10.3390/jcm11206224

41. Polinder S, Haagsma JA, van Klaveren D, Steyerberg EW, van Beeck EF. Health-related quality of life after TBI: a systematic review of study design, instruments, measurement properties, and outcome. Popul Health Metr. 2015;13:4. doi: 10.1186/s12963-015-0037-1

42. Wang B, Zeldovich M, Rauen K, Wu YJ, Covic A, Muller I, … Investigators. Longitudinal Analyses of the Reciprocity of Depression and Anxiety after Traumatic Brain Injury and Its Clinical Implications. J Clin Med. 2021;10. doi: 10.3390/jcm10235597

43. Yrysov KB, Faizullaeva GA, Mashrapov SZ. Assessing the outcomes of mild traumatic brain injury through neurocognitive testing. Science Magazine Scientific review Medical Sciences. 2021:5–9.

44. Chen W, Yao C, Li S, Huang H, Zhu Z, Chen R, … Wang G. Cognitive impairment in diffuse axonal injury patients with favorable outcome. Frontiers in Neuroscience. 2023;17. doi: 10.3389/fnins.2023.1077858

45. Scheid R, Walther K, Guthke T, Preul C, von Cramon DY. Cognitive Sequelae of Diffuse Axonal Injury. Archives of Neurology. 2006;63:418–424. doi: 10.1001/archneur.63.3.418

46. Ozen LJ, Fernandes MA. Slowing Down after a Mild Traumatic Brain Injury: A Strategy to Improve Cognitive Task Performance? Archives of Clinical Neuropsychology. 2011;27:85–100. doi: 10.1093/arclin/acr087

